# Antifungal use with and without fungal diagnoses in septic shock across U.S. hospitals, 2022-2024

**DOI:** 10.64898/2026.06.29.26355232

**Authors:** Robert J Flick, Li Yan, Anica C Law, Chad H Hochberg, Joseph Levy, Theodore J. Iwashyna, Nicholas A Bosch

## Abstract

Septic shock caused by fungal organisms is characterized by high mortality and diagnostic complexity. We used the Premier Healthcare Database to characterize antifungal use and fungal diagnoses among adults with septic shock requiring vasopressors admitted between October 2022 through July 2024. Among 12.8 million admission at 886 hospitals, 554,948 met septic shock criteria and were included for analysis. A fungal diagnosis was established in 11,405 (2.1%) of encounters; of these, 3,565 (31.3%) received intravenous antifungal therapy within one day of vasopressor initiation. In the overall cohort, antifungal therapy was initiated in 29,824 (5.5%) within one day of vasopressor initiation; of these, 3,656 (12.2%) were ultimately diagnosed with a fungal infection. In the 116 hospitals reporting microbiological data, a subgroup of 489 encounters with septic shock and culture-confirmed candidemia was identified. In this subgroup, intravenous antifungal therapy was initiated in 43.8% within one day, 63.8% within three days, and 78.9% within seven days. These findings highlight a profound decoupling between fungal diagnosis and treatment—few patients receiving antifungals were diagnosed with an infection that would be treated by these agents, while less than half of patients with septic shock and candidemia received timely treatment. Strategies for greater precision in empiric antifungal use in septic shock are needed to improve safety, stewardship, and outcomes.

## Background

Compared to bacterial organisms, septic shock from fungi is characterized by higher mortality, greater diagnostic complexity and less consensus around when empiric antimicrobials are warranted.^1–4^ Delayed antifungal administration is associated with considerable mortality among patients with fungal septic shock.^2^ We measure hospital variation in antifungal use as well as the incidence of International Classification of Diseases, tenth revision (ICD-10)-coded fungal diagnoses and culture-confirmed candidemia across the United States in patients with septic shock.

## Methods

### Description of cohort

We used the Premier Healthcare Database (PHD)which encompasses ∼25% of United States inpatient admissions.^5^ We included adults admitted between 1 October 2022 and 31 July 2024, and limited to encounters for sepsis by applying Angus sepsis algorithms (explicit sepsis diagnosis or combination of code for infection and a code for organ dysfunction) to ICD-10 diagnosis codes that were labelled as present on admission.^6,7^ We further limited to encounters with septic shock, defined as those with sepsis who were initiated on a vasopressor using hospital billing codes. Time of onset of septic shock was defined as the day of first vasopressor initiation. We characterized the cohort based on presence of at least one high-risk fungal ICD-10 discharge diagnosis code (**Table 1**). Codes describing fungal infections of an isolated non-sterile site (e.g., candida esophagitis) were excluded. We used hospital billing codes to identify use of intravenous antifungal medications within 1 (primary outcome), 3, and 7 calendar days on either side of vasopressor initiation. We used a composite of hospital death or discharge to hospice as a surrogate for 30-day all-cause mortality.^8^

**Table 1.** ICD-10 codes used to define presence of high-risk fungal infection.

| **Fungal infection** | **ICD-10 codes** |
| --- | --- |
| Aspergillosis | B44.0, B44.1, B44.7, B44.9 |
| Blastomycosis | B40.1-B40.7, B40.81, B40.89, B40.9 |
| Candidiasis | B37.6, B37.7, B37.9 |
| Coccidioidomycosis | B38.0-B38.4, B38.7, B38.89, B38.9 |
| Cryptococcosis | B45.0-B45.9 |
| Histoplasmosis | B39.0-B39.9 |
| Other specified mycoses | B48.8 |
| Paracoccidioidomycosis | B41.0-B41.9 |
| Penicillosis | B48.4 |
| Zygomycosis | B46.0-B46.9 |

ICD-10: International Classification of Diseases, tenth revision

### Other study measures

Additional variables collected for use as covariables included admission age and sex, measures of acute organ dysfunction, and Gagne combined comorbidity score (all measured at the time of admission); hospital-level factors included teaching status, census region, bed count, and urban versus rural status.^9,10^ A dummy variable for the hospital of admission was included.

### Analyses

Summary measures were stratified by presence of a high-risk fungal code using medians (interquartile range) and counts (percentages) where appropriate, with comparisons dose using Wilcoxon rank-sum tests for continuous and Chi-squared tests for categorical measures. We constructed hierarchical logistic regression models for the outcomes of antifungal use within 1, 3, and 7 days of vasopressor initiation. Included in each model were the covariables listed in the ‘other study measures’ section above. All variables were included as fixed effects except for the hospital of admission dummy variable which was included as a random effect term. From the models, we reported adjusted odds ratios for the association between each fixed effect and use of antifungals and the median odds ratio – a measure of variation in antifungal use between hospitals defined as the increased risk (in median) comparing a randomly selected hospital with lower use of antifungals to a randomly selected hospital with higher use of antifungals.^11^ We calculated Wald 95% confidence intervals for fixed effect odds ratios and used bootstrapping with 1000 iterations to calculate 95% confidence intervals for median odds ratios.

### Additional analyses

Approximately 15% of PHD hospitals contribute laboratory information. Within this subgroup we further restricted the cohort to candidemic septic shock, defined as those with a blood culture collected within 3 days either side of the day of vasopressor initiation that isolated any species of *Candida*. In a second analysis, we analyzed antifungal practices after excluding patients with oncologic (C0x-D4x) or transplant-related (Z94x) ICD-10 diagnosis codes

## Results

Of 12,814,144 adult admissions at 886 hospitals, 544,948 met septic shock criteria and 11,405 (2.1%) septic shock admissions had a diagnosed fungal infection (**Table 2**). Admissions with septic shock and a fungal diagnosis more frequently resulted in death or discharge to hospice than septic shock without fungal diagnosis (37.6% vs 32.1%, p<0.001). *Candida* (70.0%, 7,989/11,405) and *Aspergillus* (12.1%, 1,383/11,405) were the most coded fungal pathogens.

**Table 2.**
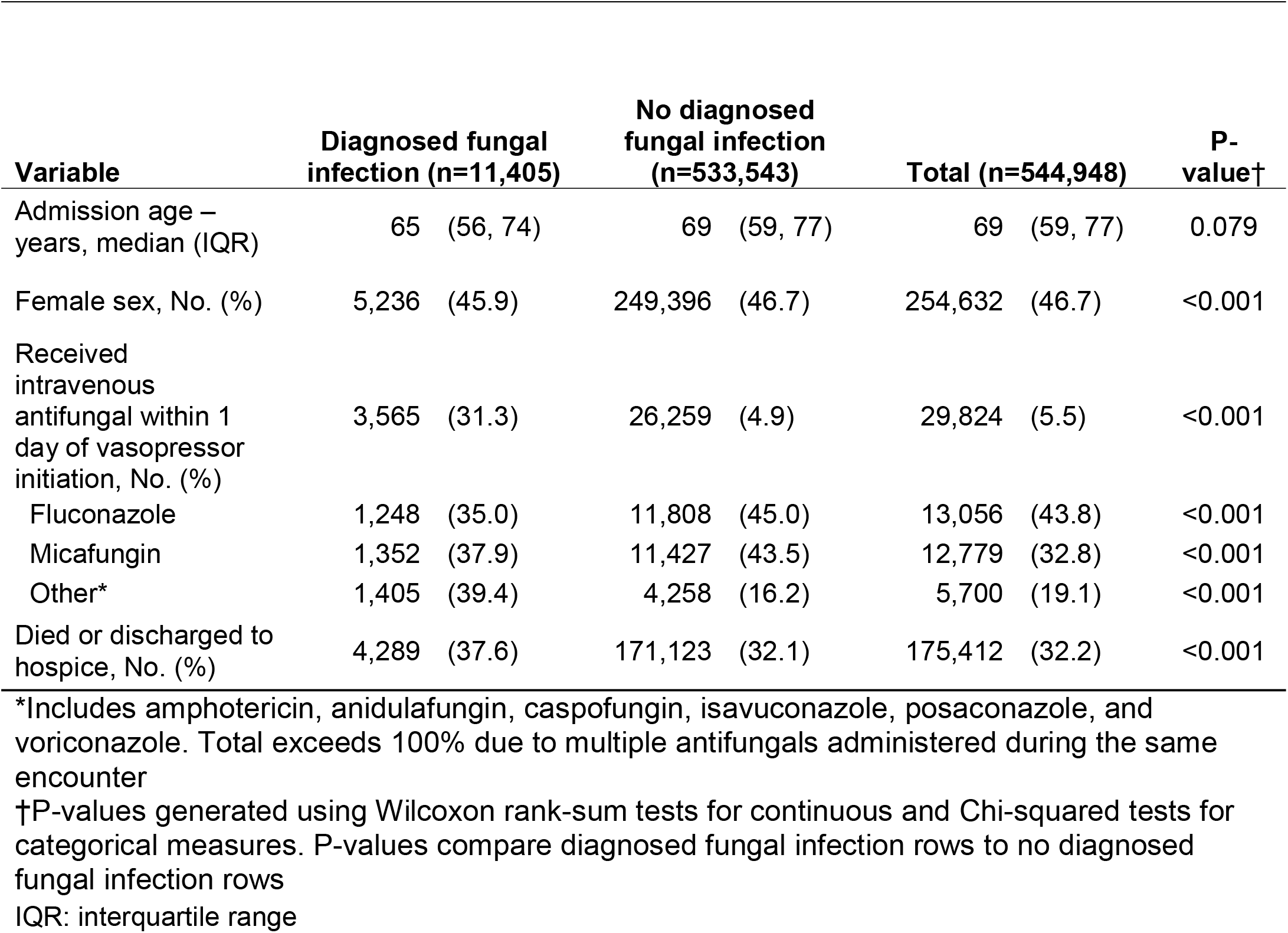
Characteristics, use of intravenous antifungals, and outcomes of hospital admission with septic shock stratified by presence of diagnosed fungal infection.

Among septic shock admissions, an intravenous antifungal was used in 29,824 admissions (5.5%) within 1 day of vasopressor initiation, 38,283 (7.0%) within 3 days, and 47,620 (8.7%) within 7 days. Fluconazole and micafungin were most common (**Table 3**).

**Table 3.** Intravenous antifungal use in patients with septic shock.

| Timing of intravenous antifungal use | Diagnosed fungal infection (n=11,405) |  | No diagnosed fungal infection (n=533,543) |  | Total (n=544,948) |  | P-value† |
| --- | --- | --- | --- | --- | --- | --- | --- |
| Within 1 day of vasopressor initiation, No. (%)* | 3,656 | (31.3) | 26,259 | (4.9) | 29,824 | (5.5) | <0.001 |
| Amphotericin | 615 | (16.8) | 542 | (2.1) | 1,157 | (3.9) | <0.001 |
| Anidulafungin | 308 | (8.4) | 1,926 | (7.3) | 2,234 | (7.5) | <0.001 |
| Caspofungin | 137 | (3.7) | 739 | (2.8) | 876 | (2.9) | <0.001 |
| Fluconazole | 1,248 | (34.1) | 11,808 | (45.0) | 13,056 | (43.8) | <0.001 |
| Isavuconazole | 99 | (2.7) | 286 | (1.1) | 385 | (1.3) | <0.001 |
| Micafungin | 1,352 | (37.0) | 11,427 | (43.5) | 12,779 | (42.8) | <0.001 |
| Posaconazole | 37 | (1.0) | 129 | (0.5) | 166 | (0.6) | <0.001 |
| Voriconazole | 246 | (6.7) | 636 | (2.4) | 882 | (3.0) | <0.001 |
| Within 3 days of vasopressor initiation, No. (%)* | 4,773 | (41.9) | 33,510 | (6.3) | 38,283 | (7.0) | <0.001 |
| Amphotericin | 777 | (16.3) | 702 | (2.1) | 1,479 | (3.9) | <0.001 |
| Anidulafungin | 437 | (9.2) | 2,556 | (7.6) | 2,993 | (7.8) | <0.001 |
| Caspofungin | 200 | (4.2) | 1,002 | (3.0) | 1,202 | (3.1) | <0.001 |
| Fluconazole | 1,713 | (35.9) | 15,209 | (45.4) | 16,922 | (44.2) | <0.001 |
| Isavuconazole | 144 | (3.0) | 382 | (1.1) | 526 | (1.4) | <0.001 |
| Micafungin | 2,026 | (42.4) | 15,010 | (44.8) | 17,036 | (44.5) | <0.001 |
| Posaconazole | 46 | (1.0) | 158 | (0.5) | 204 | (0.5) | <0.001 |
| Voriconazole | 371 | (7.8) | 841 | (2.5) | 1,212 | (3.2) | <0.001 |
| Within 7 days of vasopressor initiation, No. (%)* | 5,915 | (51.9) | 41,705 | (7.8) | 47,620 | (8.7) | <0.001 |
| Amphotericin | 936 | (15.8) | 883 | (2.1) | 1,819 | (3.8) | <0.001 |
| Anidulafungin | 567 | (9.6) | 3,337 | (8.0) | 3,904 | (8.2) | <0.001 |
| Caspofungin | 268 | (4.5) | 1,303 | (3.1) | 1,571 | (3.3) | <0.001 |
| Fluconazole | 2,243 | (37.9) | 19,015 | (45.6) | 21,258 | (44.6) | <0.001 |
| Isavuconazole | 186 | (3.1) | 480 | (1.2) | 666 | (1.4) | <0.001 |
| Micafungin | 2,667 | (45.1) | 19,344 | (46.4) | 22,011 | (46.2) | <0.001 |
| Posaconazole | 58 | (1.0) | 184 | (0.4) | 242 | (0.5) | <0.001 |
| Voriconazole | 556 | (9.4) | 1,115 | (2.7) | 1,671 | (3.5) | <0.001 |

\*Total exceeds 100% due to multiple antifungals administered during the same encounter.
†P-values generated using Chi-squared comparing high risk fungal infection code present to high risk fungal infection code absent columns.

Among septic shock admissions with a fungal diagnosis, 31.3% (3,565/11,405) received an intravenous antifungal within 1 day of vasopressor initiation. The majority of intravenous antifungal use within 1 day was in admissions where a fungal infection was not documented in diagnostic codes (88.0%, 26,259/29,824). This pattern persisted (87.5%, 18,473/21,106) even after excluding admissions with transplant or oncologic diagnoses, where prophylactic antifungal use may be more common ^12^.

A subgroup of 116 hospitals (14%) reported blood culture results to the database. A total of 489 admissions to these hospitals had blood cultures drawn at the time of vasopressor initiation that subsequently confirmed candidemia. Intravenous antifungal therapy was initiated in 43.8% (214) of these cases within 1 day, 63.8% (312) within 3 days, and 78.9% (386) within 7 days of vasopressor initiation. Of these 489 candidemia cases, blood cultures were finalized on the day of death or later in 32.1% (157) of cases.

In multivariable analysis for receipt of intravenous antifungals within 1 day of vasopressor initiation, patient-level factors such as additional vasopressor use (odds ratio [OR] = 1.45 for each additional vasopressor, 95% confidence interval [CI] 1.43-1.47), acute renal dysfunction (OR=1.38, 95%CI 1.34-1.42), and acute respiratory dysfunction (OR=1.26, 95%CI 1.23-1.29) were of similar magnitude as the variation in practice between hospitals (median odds ratio 1.55, 95%CI: 1.53 – 1.58; **Figure 1**).

**Figure 1.**
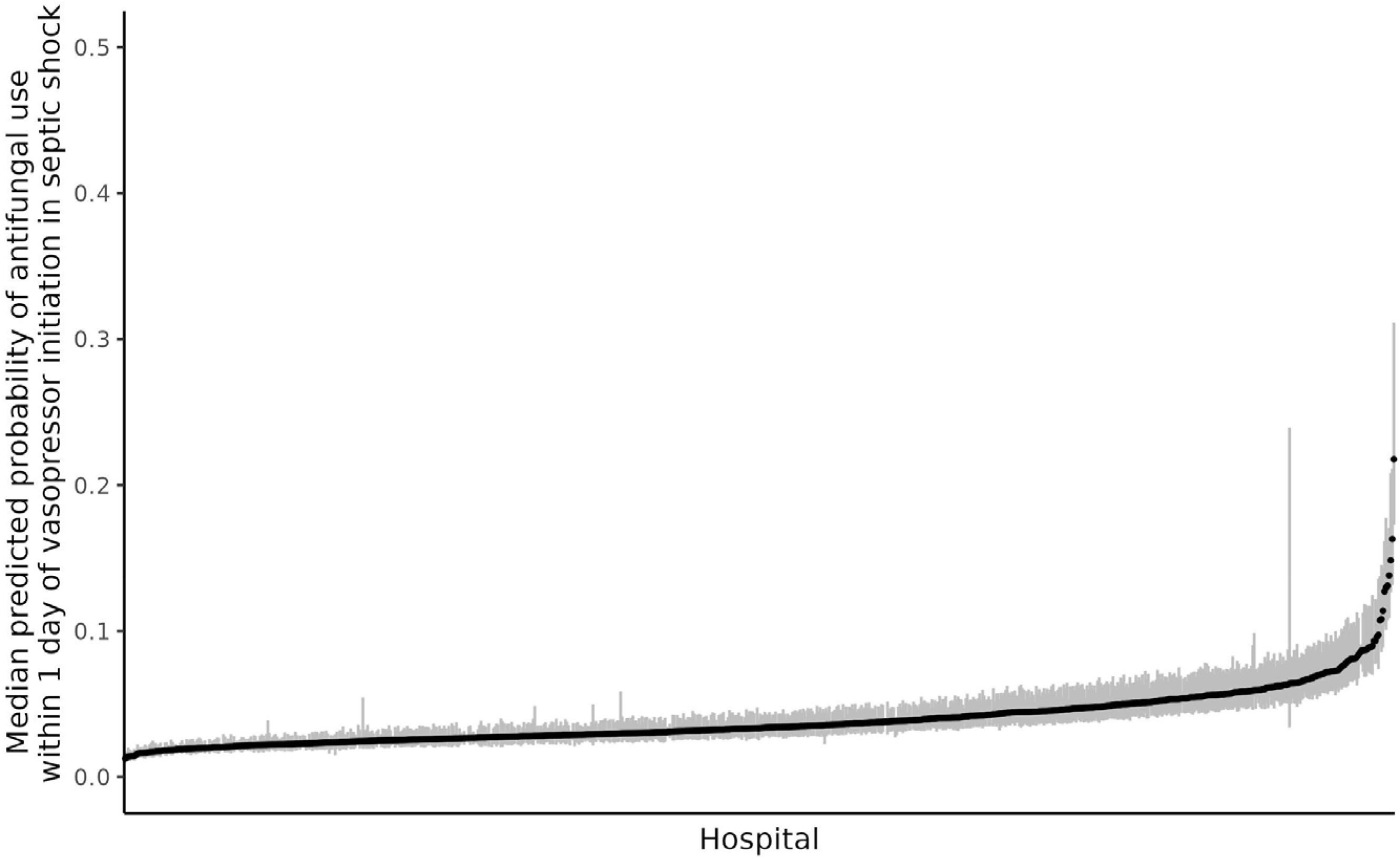
Median predicted probabilities of antifungal use within 1 day of vasopressor initiation per hospital among septic shock encounters. Shown are medians (black dots) and interquartile ranges (grey lines) predicted probabilities from the hierarchical logistic regression models for antifungal use within 1 day of vasopressor initiation per hospital.

## Discussion

In this national sample of contemporary patients with septic shock, many patients with diagnosed fungal infections—including culture-confirmed candidemia—did not receive timely intravenous antifungal therapy. Although antifungal use was more frequent in those with a fungal diagnosis, most antifungal use occurred in those without a fungal diagnosis. There was substantial hospital variation in the proportion of early intravenous antifungal use.

These results raise the possibility that more tailored use of antifungals could simultaneously improve safety, stewardship, and outcomes. Important limitations of this work include reliance on blood culture confirmation of candidemia or on administrative codes, neither of which are perfect for measuring septic shock attributable to a fungal pathogen.

## Data Availability

Data accessed under a commercial contract and cannot be shared

## References

1. Martin-Loeches I, Cornely OA, Denning DW, et al. Invasive candidiasis in intensive care medicine: shaping the future of diagnosis and therapy. Intensive Care Med. 2025;51(11):2065–2078. doi:10.1007/s00134-025-08151-1

2. Kollef M, Micek S, Hampton N, Doherty JA, Kumar A. Septic shock attributed to Candida infection: importance of empiric therapy and source control. Clin Infect Dis. 2012;54(12):1739–1746. doi:10.1093/cid/cis305

3. Stempel JM, Farmakiotis D, Tarrand JJ, Kontoyiannis DP. Time-to-reporting of blood culture positivity and central venous catheter-associated Candida glabrata fungemia in cancer patients. Diagn Microbiol Infect Dis. 2016;85(3):391–393. doi:10.1016/j.diagmicrobio.2016.04.001

4. Evans L, Rhodes A, Alhazzani W, et al. Surviving Sepsis Campaign: International Guidelines for Management of Sepsis and Septic Shock 2021. Crit Care Med. 2021;49(11):e1063–e1143. doi:10.1097/CCM.0000000000005337

5. Premier Applied Sciences, Premier Inc. Premier Healthcare Database: Data That Informs and Performs (White Paper). 2025. https://offers.premierinc.com/Premier-Healthcare-Database-Download.html

6. Iwashyna TJ, Odden A, Rohde J, et al. Identifying patients with severe sepsis using administrative claims: patient-level validation of the angus implementation of the international consensus conference definition of severe sepsis. Med Care. 2014;52(6):e39–43. doi:10.1097/MLR.0b013e318268ac86

7. Bosch NA, Law AC, Rucci JM, Peterson D, Walkey AJ. Predictive Validity of the Sequential Organ Failure Assessment Score versus Claims-based Scores among Critically Ill Patients. Ann Am Thorac Soc. 2022;19(6):1072–1076. doi:10.1513/AnnalsATS.202111-1251RL

8. Prescott HC, Heath M, Jayaprakash N, et al. Concordance of 30-Day Mortality and In-Hospital Mortality or Hospice Discharge After Sepsis. JAMA. 2025;333(19):1724–1726. doi:10.1001/jama.2025.2526

9. Sun JW, Rogers JR, Her Q, et al. Adaptation and Validation of the Combined Comorbidity Score for ICD-10-CM. Med Care. 2017;55(12):1046–1051. doi:10.1097/MLR.0000000000000824

10. Gagne JJ, Glynn RJ, Avorn J, Levin R, Schneeweiss S. A combined comorbidity score predicted mortality in elderly patients better than existing scores. J Clin Epidemiol. 2011;64(7):749–759. doi:10.1016/j.jclinepi.2010.10.004

11. Merlo J, Chaix B, Ohlsson H, et al. A brief conceptual tutorial of multilevel analysis in social epidemiology: using measures of clustering in multilevel logistic regression to investigate contextual phenomena. J Epidemiol Community Health. 2006;60(4):290–297. doi:10.1136/jech.2004.029454

12. Taplitz RA, Kennedy EB, Bow EJ, et al. Antimicrobial Prophylaxis for Adult Patients With Cancer-Related Immunosuppression: ASCO and IDSA Clinical Practice Guideline Update. J Clin Oncol Off J Am Soc Clin Oncol. 2018;36(30):3043–3054. doi:10.1200/JCO.18.00374

